# Pre-TAS Evaluation of mass drug administration effectiveness on lymphatic filariasis elimination and study of Immunological response in elephantiasis patients in Osun State, Nigeria

**DOI:** 10.64898/2026.09.14.26363010

**Authors:** Samuel Chibueze Nzekwe, Babarinde Samuel Olufolarin, Abiodun Lateef Boladale, Adetoun Elizabeth Morakinyo, Uzoma Emeka, Tolulope Abolaji Adeyemo, Damilale Oluwafemi Kehinde, Adeyemi Temitayo Adeyemo, Marvelous Olasunmbo Ogundinmu, David Anuoluwapo Oyebamiji, Elijah Oluwatosin Olopade, Oladunni Opeyemi, Anyaele Okoeghua Okorie, Musibau Olaniyi Adejumobi, Alexander Bababunmi Odaibo, Godwin Anyim, Adekunle Isiaka, Adie Hilary, Ademola Oluboade Ayeleso

## Abstract

The need to meet the World Health Organization 2030 target for elimination of lymphatic Filariasis necessitates various activities such as mass administration of medicines (MAM), pre-transmission assessment surveys (pre-TAS)/ epidemiological monitoring survey (EMS) and transmission assessment survey. However for this study, pre-TAS/EMS was conducted to validate elimination milestones and elucidate the lingering immunological consequences of chronic infection. This study conducted a comprehensive evaluation across twelve Local Government Areas (LGAs) in Osun State, screening,7388 individuals for circulating filarial antigen (CFA) using Abbott Filarial Test Strips (FTS), while simultaneously investigating immune modulation in patients with clinical elephantiasis through detailed haematological profiling. Results indicated that the prevalence of the disease at sentinel and spot check sites have been lowered below threshold levels thereby qualifying for a first transmission assessment survey (TAS1) , evidenced by a 0% antigenemia rate across all surveyed LGAs, a figure significantly below the World Health Organization’s <2% threshold. . The average percentage of individual participation in MDA/MAM programs was found to be 56.09%, whereby, Ola-Oluwa LGA recorded the highest at 84.70% while the list MDA program participation was found at Ilesha East with 18.97%, highlighting a potential vulnerability in vector control compliance. Furthermore, haematological assessments of chronic cases revealed a distinct state of immune suppression, characterized by significant reductions in total white blood cell and lymphocyte counts, alongside persistent monocytosis and eosinopenia. These findings suggest that while Osun State has successfully transitioned into a post-transmission phase, the lingering physiological impacts on the chronically ill and the moderate rates of preventive practice necessitate sustained surveillance and enhanced public health interventions to ensure permanent disease eradication

**Author Summary:** Lymphatic filariasis (LF) is a debilitating neglected tropical disease primarily caused by *Wuchereria bancrofti* and transmitted by mosquitoes, leading to chronic physical deformities such as elephantiasis. Although global infections have decreased, LF remains a critical public health concern across Africa. Nigeria reports a national prevalence rate of 11.18%, with the South West zone contributing 1.26%. Prior regional surveys in Osun State documented a 1.7% circulating filarial antigen (CFA) prevalence, highlighting the necessity of systematic mass drug administration (MDA) and continuous post-intervention monitoring.

To assess transmission interruption and host immunological status, this study screened 7,388 individuals across 12 Local Government Areas (LGAs) in Osun State using Abbott Filarial Test Strips (FTS). Results revealed a 0% CFA prevalence across all 24 sentinel and spot-check sites, successfully meeting the World Health Organization threshold (<2%) to qualify the region for its first Transmission Assessment Survey (TAS1). Survey metrics demonstrated an average MDA participation rate of 56.09%, whereas long-lasting insecticidal net (LLIN) compliance averaged significantly lower at 41.09%. These findings confirm the interruption of active LF transmission while emphasizing the necessity of sustained post-MDA surveillance to prevent disease resurgence.

## Introduction

Lymphatic filariasis (LF) is a debilitating neglected tropical disease caused by parasitic worms, primarily *Wuchereria bancrofti*, which is responsible for about 90% of cases, while *Brugia malayi* and *Brugia timori* account for the remaining infections. The parasites are transmitted through mosquito vectors and can lead to progressive disability and physical deformity, most notably elephantiasis [1]. History accounts that LF is common in Central and West Africa, Nile Delta, Thailand, Pakistan, India, Korea, Japan, Philippines, in a geospatial analysis report. An estimated global population of 199 million people was infected with lymphatic filariasis, including 3.1 million people in America and 107 million in South East Asia by 2000 but experienced a sharp decline by 2018 except in Africa and South East Asia where local elimination are yet to reach the threshold [2].

In Nigeria, the review work of Waje et al., [3] noted a national prevalence rate of 11.18% of lymphatic filariasis. The results revealed a positive case of 1,146 (63.00%) for microfilaria and 673 (37.00%) were positive for Circulating Filarial Antigen (CFA). This national prevalence rate is composed of contributions from various geopolitical zones, including the North Central and North East at 4.52%, the South-South and South East at 3.81%, the North West at 1.59%, and the South West at 1.26%. It was noted in their review that 566 of the positive subjects (31.12%) presented various clinical symptoms. The National Lymphatic Filariasis Elimination Programme (NLFEP) which was established in 1997 has the mandate to eliminate LF as a public health problem in Nigeria. Presently, all the 583 endemic LGAs have conducted at least one round of mass administration of medicine (MAM). Out of the 583 LGAs that have conducted MAM, 348 LGAs have stopped treatment having passed first Transmission Assessment Survey (TAS1) [4]. Lymphatic filariasis case report from North West Nigeria, by Branta et al., (2018) specifically through micro-stratification overlaps mapping (MOM) and surveillance literature, reported a 10.00% CFA prevalence and 0.3% microfilaria rate for Kaduna State. Similarly, Katsina State reported a 7.30% CFA prevalence via Immunochromatographic Test (ICT) and 0.90% via microscopy during baseline outcome mapping according to Eneanya et al. [5]. In Zamfara State, a study of six communities in the Talata Mafara Local Government Area revealed a high seroprevalence rate of 37.80% CFA by Rapid Diagnostic Test (RDT), attributing the high figures to poor sanitation, proximity to water bodies, and a general lack of awareness [6].

In South West Nigeria, research in the Ado-Odo Ota and Abeokuta South LGAs of Ogun State reported microfilaria prevalence rates of 4.00% and 2.40% respectively. These findings were linked to inadequate awareness regarding the causes of the disease. Furthermore, Sowo Village in Abeokuta reported a 17.00% microfilaria prevalence alongside a 2.20% incidence of hydrocele and elephantiasis, occurring in an environment characterized by dense vegetation, lack of drainage, and the presence of *Aedes*, *Culex*, and *Anopheles* mosquito vectors [7]. In Osun State, an immunochromatographic card test across ten selected communities in five LGAs revealed a 1.70% CFA prevalence, with documented cases of hydrocele and the discovery of *Anopheles gambiae* mosquitoes infected with *Wuchereria bancrofti* [3].

Implementation of Mass Drug Administration (MDA) in Osun State has shown promising results in reducing transmission, with numerous endemic areas now meeting the criteria to scale down interventions following years of effective coverage. However, the report from Federal Ministry of Health (2024) emphasizes that maintaining these gains requires rigorous post-MDA surveillance to promptly detect any resurgence and ensure the momentum toward elimination is sustained [4].

The complexity of LF is further compounded by the parasite’s sophisticated modulation of the host immune responses, which enables long-term survival [8]. This dynamic interaction requires the parasite to undergo metabolic switches to adapt to drastically different environments, from the mosquito to the human host [9]. In humans, they evade both innate and adaptive immune mechanisms. The innate immune system, involving natural killer (NK) cells, eosinophils, and macrophages, is often subverted. Research has shown that infective larvae can rapidly activate NK cells, followed by apoptosis, thereby weakening early host defences [10]. Similarly, immune mediators such as IL-5 and eosinophils, typically involved in parasite clearance, may sometimes signal accelerated larval growth [10]. Adaptive immune response also plays a crucial role in controlling filarial infections. Antigen presentation by macrophage activates CD4+ T cells and promotes cytokine secretion (IL-3, IL-4, IL-5, IL-9) leading to mast cell activation eosinophil recruitment, and B-cell-mediated antibody production [11,12]. Specific antibody profiles have diagnostic and functional significance. For example, high levels of IgG4, are often indicative of a *W. bancrofti* infection, while IgG1 may confer protection against *B. malayi* [13]. Effector mechanisms, including antibody-dependent cellular cytotoxicity and complement-mediated lysis, further contribute to parasite clearance [14,15]. Additionally, evaluating haematological profiles of LF patients provides insights into the disease’s impact on immune function and support future morbidity management and elimination strategies, including the development of targeted anthelmintic therapies. Therefore, this study aimed to determine the prevalence of lymphatic filariasis across 12 LGAs of Osun State, Nigeria, and assess the haematological profiles and immune cells mobilisation in patients presenting with elephantiasis-related complications. This study assessed the prevalence of lymphatic filariasis in 12 LGAs (Orolu, Osogbo, Boluwaduro,

Ifedayo, Iwo, Ola-Oluwa, Aiyedade, Isokan, Ife North, Ife South, Ede South and Ilesa East) of Osun State, Nigeria, and examined haematological profiles of immune cells, including white blood cell (WBC), lymphocyte, monocyte, and eosinophil counts in individuals with clinical manifestations of lymphatic filariasis. The results provide compelling evidence that active transmission of LF in the surveyed LGAs has been successfully interrupted, as demonstrated by the 0% prevalence rate recorded across all sampled individuals using the Abbott Filarial Test Strip. This indicates that Osun State, Nigeria has attained the post-transmission phase of LF elimination, demonstrating the success of mass drug administration (MDA), vector control, and community-based health interventions.

Nonetheless, the haematological analyses of previously diagnosed LF patients revealed lingering immune alterations, characterised by variable levels of WBCs, lymphocytes, neutrophils, monocytes, eosinophils and the absence of basophils. This work showed ongoing immune modulation even after parasite clearance, reinforcing the necessity of continuous care for chronic cases through morbidity management and disability prevention (MMDP) programmes, as recommended by the WHO.

## Methods

### Informed Consent Statement

All participants involved were duly informed and filled out participation forms.

### Pre-survey community mobilization and sensitization

Pre-survey visit was carried out in collaboration with the State Ministry of Health (SMOH) team, UNICEF, AHF and LGA NTDs coordinators of the 12 LGAs. During the visit, the community leaders and members were sensitized on the planned survey.

### Study Design and sample area

Pre-Transmission Assessments were carried out in 12 Local Government Areas (LGAs) of Osun State, Nigeria (Aiyedade, Boluwaduro, Ede South, Ife North, Ife South, Ifedayo, Ilesha East, Isokan, Iwo, Ola Oluwa, Orolu, and Oshogbo) from September 17–25, 2024, by the Federal Ministry of Health (FMOH) in collaboration with the Department of Public Health, Ministry of Health, Osun State, Nigeria, and the Department of Biochemistry, Adeleke University, Ede, Osun State, with support from the United Nations Children’s Fund (UNICEF) and Amen Health & Empowerment Foundation (AHF). Twenty-four (24) sites (i.e., 12 sentinel sites and 12 spot-check sites) were selected for the survey across the 12 LGAs of the State.

A minimum of 300 valid blood Samples were collected per site in the LGA. The target groups were community members ages ≥5 years irrespective of gender qualified to participate in the survey voluntarily. The data forms were used for the survey including community, individuals and summary forms to obtain information on the use of long lasting insecticidal nets (LLINs) and participation in MDA. A total of seven thousand three hundred and eighty-eight (7388) individuals across the 12 LGAs, and a sample size of not less than 600 participants in each local government, according to the WHO standard as shown in Figure 1 below.

**Fig 1:**
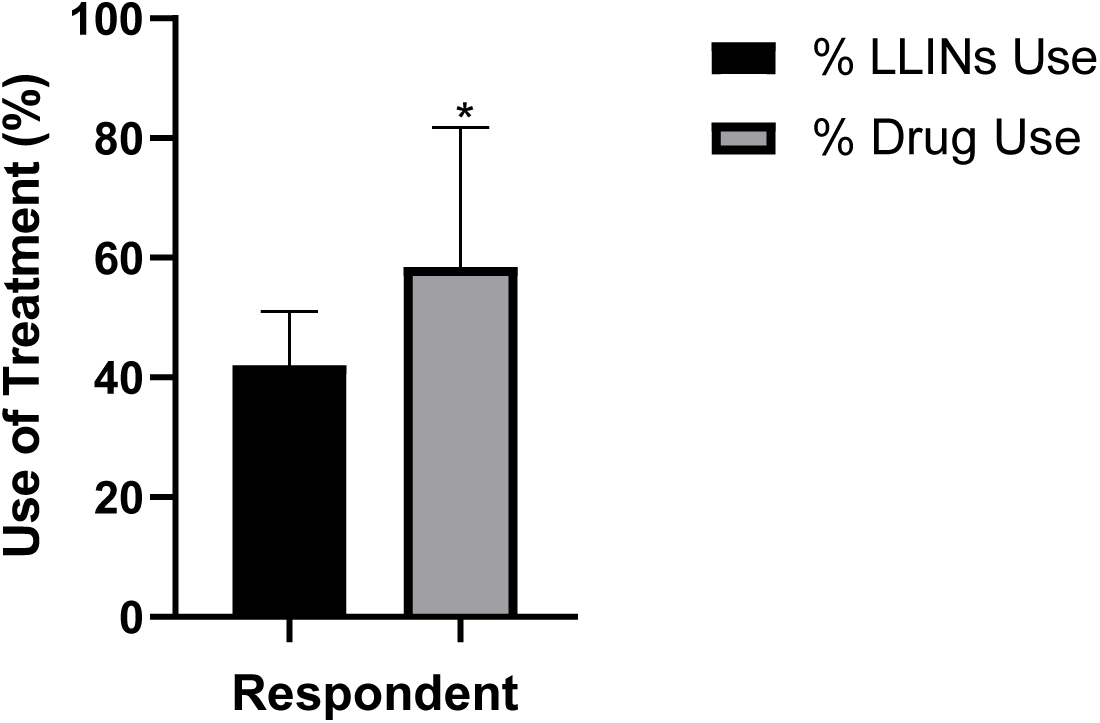
Comparison of the percentage of respondents using long-lasting insecticidal nets (LLINs) and participating in mass drug administration (MDA) for lymphatic filariasis at sentinel sites. Values were expressed as median (interquartile range) using nonparametric Wilcoxon test. (*) was statistically (W = 52, p = 0.0425) higher than (%LLINs Use).

## Methods used to carryout Pre-Transmission Assessment Survey

### Pre-survey training

A two-days training was held at the State capital. The training was held at Leisure Spring Hotel Oshogbo, Osun State from 17th to 18th September, 2024. The training content included: Overview of LF Elimination activities in the State, LF Pre-TAS guidelines, logistics, data management reporting format (hard copy of community, Individual and summary forms) interview techniques, effective usage of FTS kits etc.

### Sample collection and test procedure

Data were collected from age group ≥5 years that are residents of the selected communities. Participation in the survey was voluntary. Verbal consent was obtained from adults and assent from parent(s) or guardian for children. Blood samples (75µl) from the consenting participants were collected via finger-prick and then emptied by drops using the calibrated capillary tubes onto the sample pad of the Abbott Filarial Test Strip (FTS), an immunochromatographic test (ICT) strip used for detecting circulating filarial antigen [16]. The test result was read after 10 minutes. In a positive test, the control and test lines appear to indicate the presence of lymphatic filarial antigen with a positive result (presence of filarial antigen) indicated by the appearance of both control and test lines, while the appearance of only the control line indicated a negative result.

### Immunological Biomarkers assessment

In addition, blood samples were collected from elephantiasis patients, in Ethylene-diamine-tetra-acetic acid-containing anticoagulant tubes (50uL) for haematological immune assays. Elephantiasis disease condition was confirmed on the patients by medical consultants at University of Osun State Teaching hospital and through physical manifestation lower limb lymphangitis (elephantiasis). The control samples patients were tested with Abbott Filarial Test Strip (FTS) to confirm the absence of filarial antigen. The blood samples were diluted with a cell pack solution in the White Blood Cell (WBC) counting chamber. A fixed volume of Stromatolyser-W solution (1 volume of Stromatolyser-WH to 2 volumes of cell pack) was added to obtain a final dilution of 1:500. The cells were subsequently counted by the Direct Counted Method (DCM). Haemoglobin released during RBC lysis was converted to the red methaemoglobin and read photometrically at 555 nm.

### Statistical Analysis

All collected field data from the 12 LGAs of the state were verified by examining the field hard copies and analyzed using excel sheet. Health Mapper was used to confirm the survey sites’ coordinates. To avoid bias, completed field data forms were interchanged among team leaders, and issues highlighted were reconciled by the team leaders who collected the data. Data analysis of blood samples from elephantiasis patients and controls was performed using two-way ANOVA at p < 0.05 level of significance. Values were expressed as Mean ± SEM (n = 6) using the GraphPad Instat (GPIS) package, version 5. The positive results were compared against the mean of randomly selected outpatients in the UNIOSUN teaching hospital using the Bonferroni post hoc test.

## Results

### Participants’ response on usage of Long-Lasting Insecticidal Nets (LLINs) and Participation in Mass Drug Administration (MDA) in sentinel sites

The percentage of respondents using long-lasting insecticidal nets (LLINs) and participating in mass drug administration (MDA) for lymphatic filariasis at sentinel sites was presented in Figure 1. A Wilcoxon matched-pairs rank test showed that there was a statistically significant (W = 52, p = 0.0425) difference between the %Drug Use and %LLINs Use. This shows that LLIN usage was significantly lower than MDA drug uptake across the sentinel sites studied. The percentage of respondents using long-lasting insecticidal nets (LLINs) ranged from 28.38 to 60.32% across the 12 sites, with a median of 40.43% (IQR: 36.39 – 48.11%; mean = 42.03 ± 9.03%). In contrast, participation in mass drug administration (MDA) ranged from 18.97 to 81.43%, with a median of 64.84% (IQR: 36.21 – 78.69%; mean = 58.46 ± 23.38%). The median within-site difference (%Drug use − %LLIN use) was 25.09 percentage points (IQR: −10.27 to 35.51%). This shows that, at most sites, a considerably higher proportion of respondents participated in MDA drug distribution than using LLINs.

### Participants’ response on usage of Long-Lasting Insecticidal Nets (LLINs) and Participation in Mass Drug Administration (MDA) in spot check sites

The percentage of respondents that used LLINs and participated in MDA for LF at spot check sites (SC) was presented in Figure 2. The mean percentage of respondents using LLINs across sentinel sites was 40.22 ± 7.487% (95% CI: 35.46–44.98%), while the mean percentage participating in MDA drug distribution was 53.56 ± 18.20% (95% CI: 41.99–65.12%). A paired t-test revealed that %Drug use was significantly higher than %LLIN use across the 12 spot sites, with a mean difference of 13.34 percentage points (95% CI: 2.475– 24.20; t = 2.703, df = 11, p = 0.0206). The correlation coefficient between paired LLIN and drug use values was weak and non-significant (r = 0.3490, one-tailed p = 0.1331). This shows that the pairing by site did not significantly improve the precision of the estimate.

**Fig 2:**
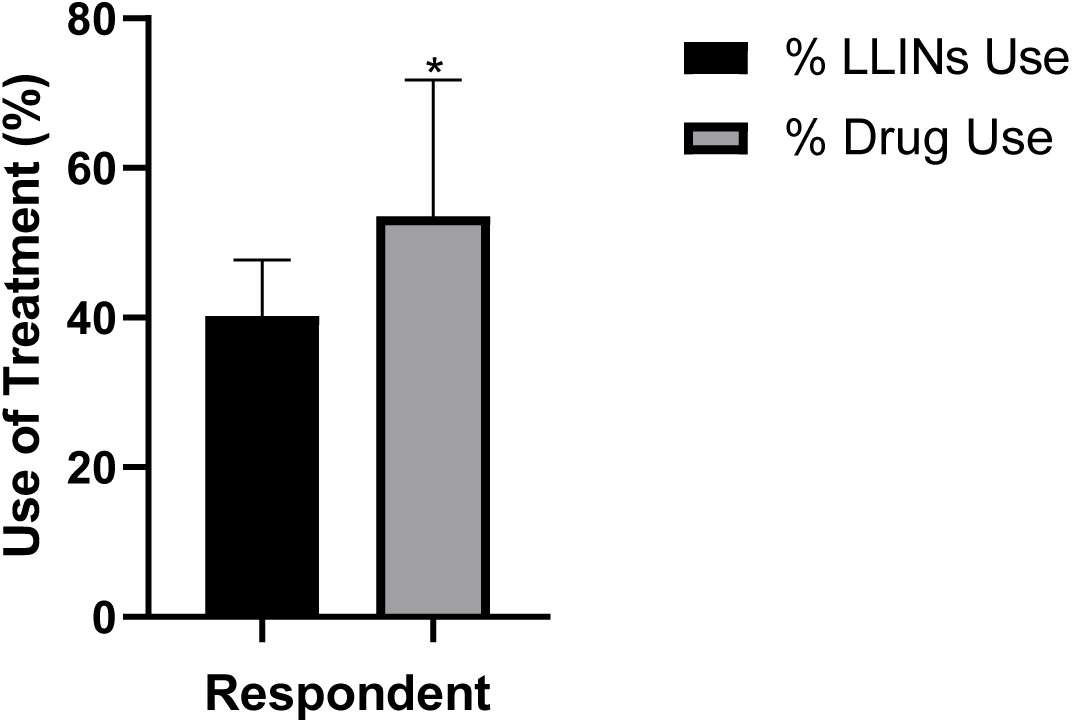
Percentage of respondents that used LLINs and participated in MDA for LF at spot check sites (SC). Values were expressed as Mean ± SD. Comparison between %LLIN use and %Drug use across sites was performed using a paired t-test, with p < 0.05 considered statistically significant. (*) is significantly (p = 0.0206) higher than %LLINs use.

### Lymphatic Filariasis Pre-Transmission Assessment Survey (Pre-TAS) in 12 LGAs of Osun State in sentinel sites

The level of participation of male and female in LF Pre-TAS in 12 LGAs of Osun State at sentinel site (SS) was presented in Figure 3. Across the 12 sites, the mean number of male respondents was 120.5 ± 18.07 (95% CI: 109.0–132.0), while the mean number of female respondents was 186.7 ± 19.50 (95% CI: 174.3–199.1). A paired t-test showed that the number of female respondents was significantly higher than male respondents at each site, with a mean difference of 66.17 (95% CI: 42.69–89.64; t = 6.203, df = 11, p < 0.0001). The correlation coefficient between paired male and female counts was strong and negative (r = −0.9345, p < 0.0001), and the pairing was found to be significantly effective. This means that as the number of male respondents increased at a site, the number of female respondents tended to decrease correspondingly.

**Fig 3:**
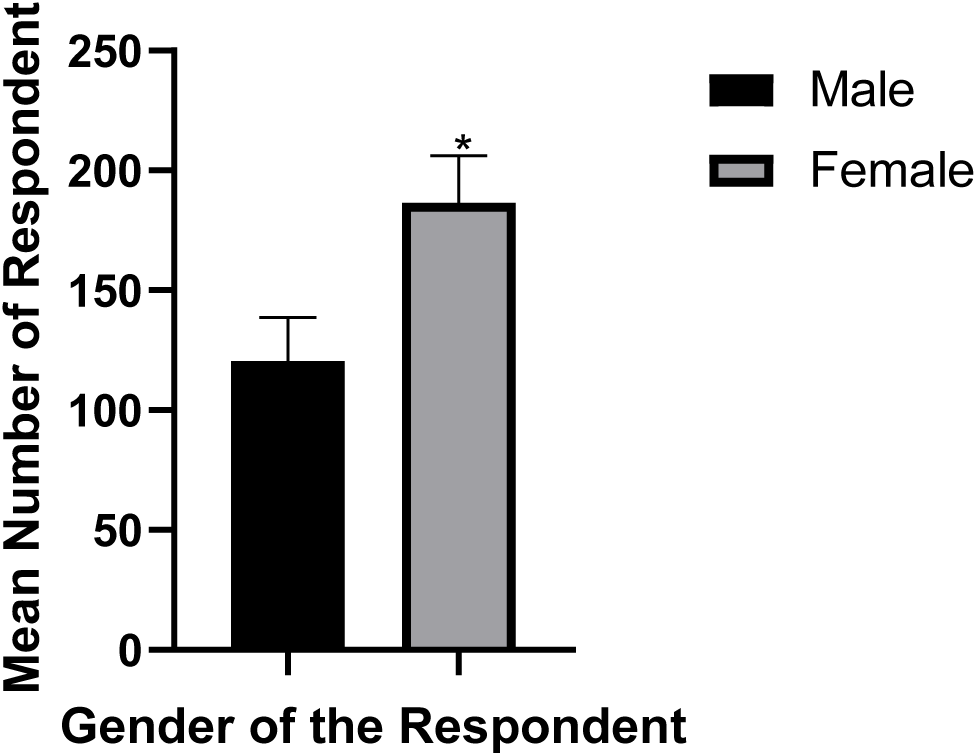
Level of participation of male and female in LF Pre-TAS in 12 LGAs of Osun State at sentinel site (SS). Values were expressed as Mean ± SD. Comparison between male and female across sentinel sites was performed using a paired t-test, with p < 0.05 considered statistically significant. (*) is highly significantly (p = 0.0206) higher than male.

### Lymphatic Filariasis Pre-Transmission Assessment Survey (Pre-TAS) in 12 LGAs of Osun State in spot check sites

The level of participation of male and female in LF Pre-TAS in 12 LGAs of Osun State at spot check site (SC) was presented in Figure 4. Across the 12 sentinel sites, the mean number of male respondents was 118.5 ± 19.61 (95% CI: 106.0 – 131.0), while the mean number of female respondents was 189.0 ± 21.67 (95% CI: 175.2–202.8). A paired t-test showed that female respondents significantly outnumbered male respondents at each site, with a mean difference of 70.50 (95% CI: 44.55–96.45; t = 5.979, df = 11, p < 0.0001). The correlation coefficient between paired male and female counts was strong and negative (r = −0.9583, p < 0.0001), and the pairing was found to be significantly effective.

**Fig 4:**
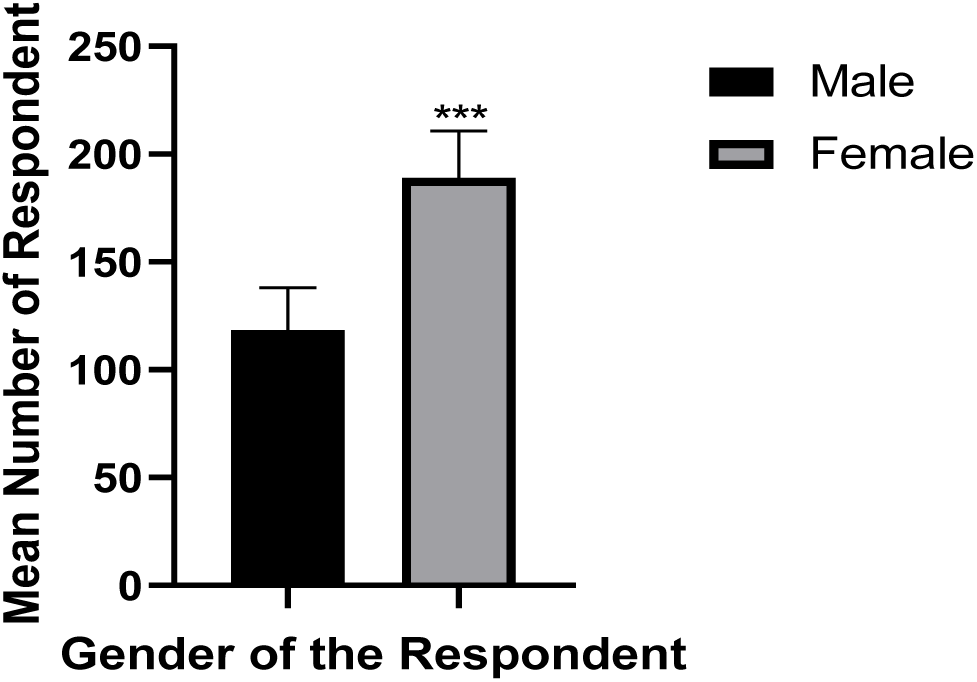
Level of participation of male and female in LF Pre-TAS in 12 LGAs of Osun State at spot check site (SC). Values were expressed as Mean ± SD. Comparison between male and female across spot check sites was performed using a paired t-test, with p < 0.05 considered statistically significant. (****) is highly significantly (p = <0.0001) higher than male.

### Haematological Assessment of Immune Cell Expression in the Blood of LF Patients and Control Samples

The haematological profile of LF complicated individuals were assessed as shown in Figure 5. The results evaluation revealed statistically significant variations (p < 0.05) in white blood cell (WBC), lymphocyte, monocyte, and eosinophil counts in LF patients compared to controls. LF Patients A and B exhibited a marked decrease in white blood cells (WBCs), lymphocytes, neutrophils, and eosinophils, but an increase in monocyte levels. LF Patients C and D demonstrated a significantly increased counts of WBCs, lymphocytes, neutrophils, and monocytes. Patient E showed elevated levels of WBCs, lymphocytes, and monocytes, but a reduced neutrophil and eosinophil counts.

**Fig 5:**
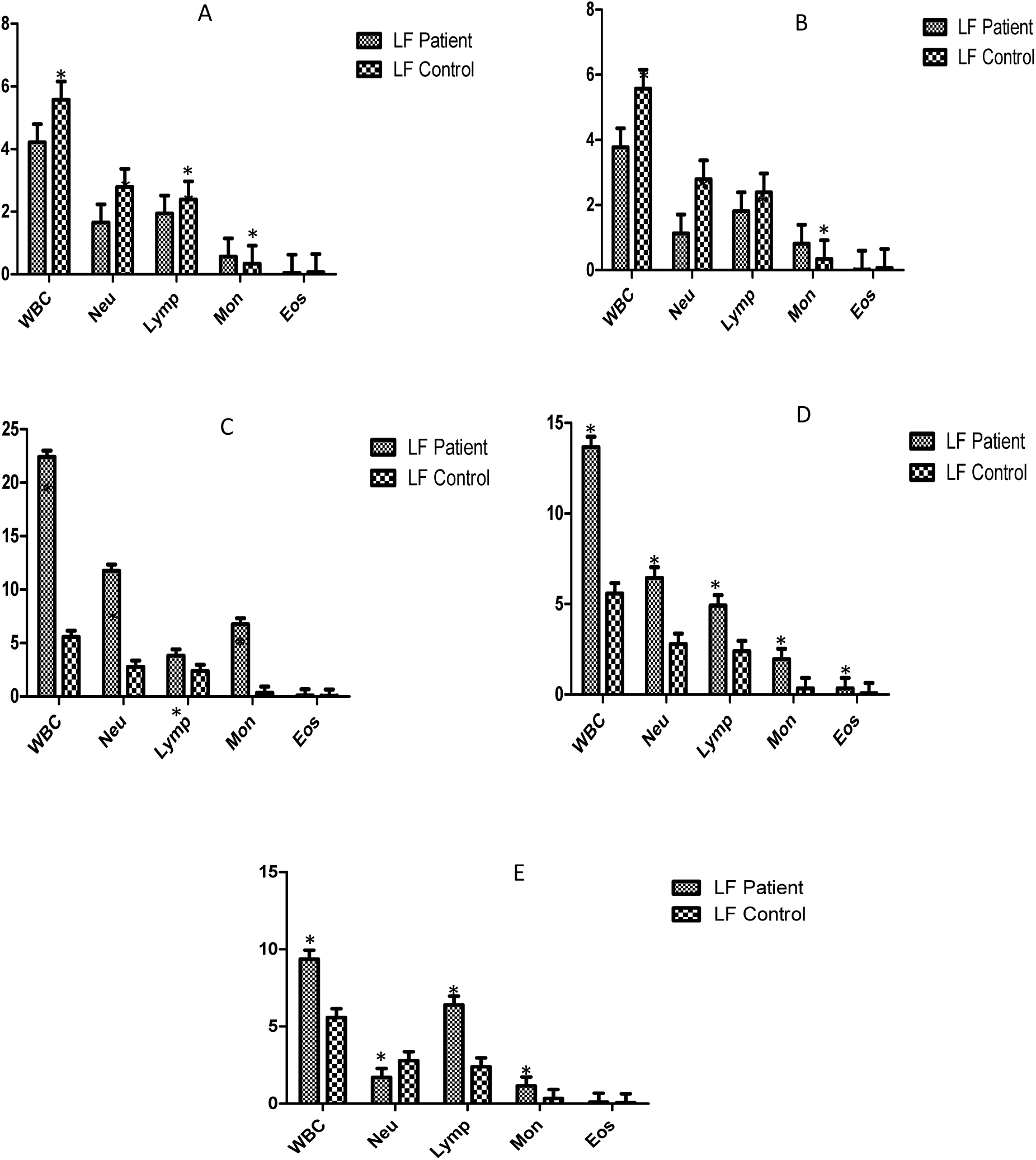
Haematological Assessment of Immune Cell Concentrations in LF Patients and Controls. A, B, C, D and E represent the results of haematological assays in blood samples of Patients A, B, C, D, and E compared to healthy control samples. White blood cells (WBC), Neutrocytes (Neu), Lymphocytes (Lymp), Monocytes (Mon) and Eosinophils (Eos) were the Immune Cells analysed. (*) indicate a significant difference between the immune cells of LF patients and the control group. Statistical significance level was set at p<0.05.

## Discussion

Lymphatic filariasis (LF) remains a neglected tropical disease affecting millions globally, despite extensive elimination efforts. Lymphatic filariasis (LF) has been defined by the complex interplay between the parasite and the host immune system, which results in a spectrum of clinical outcomes ranging from asymptomatic infection to severe lymphatic pathology such as lymphedema and elephantiasis, among other symptoms [17]. While the interruption of transmission is a key goal, the long-term immunological effects on individuals with existing complications like elephantiasis remain poorly understood. This study assessed the current transmission status of LF and the haematological profiles of infected individuals across selected Local Government Areas (LGAs) in Osun State, Nigeria. In this pre-TAS study, a total of 7,388 individuals from 12 LGAs were screened for circulating filarial antigen (CFA) using Abbott Filarial Test Strips (FTS). Additionally, haematological parameters, including white blood cells (WBCs) such as lymphocytes, monocytes, neutrophils, and eosinophils, were evaluated in five previously diagnosed LF patients to determine immune cell modulation associated with chronic infection. The pre-transmission assessment survey (pre-TAS) yielded a 0% antigenemia rate across all 12 LGAs, which falls within the WHO threshold (< 2%) for the cessation of mass drug administration (MDA). The pre-transmission assessment survey (pre-TAS) revealed a 0% antigenemia rate across all LGAs, which is below the World Health Organization (WHO) threshold of <2% required for stopping mass drug administration (MDA), suggesting interruption of transmission in the study areas. Preventive practices showed moderate or below LLINs usage with average of 41.09% of participants using long-lasting insecticide-treated nets (LLINs) in all the LGAs studied. The highest percentage was recorded at Ola-Oluwa LGA with 60.32% and the lowest percentage LLIN usage compliance at Oshogbo LGA with 27.35%. Greater percentage of the LGAs reported more than average participation in MDA programme. Greater number of the LGAs studied showed higher percentage in drug usage or MDA participation than the average. The areas such as Oshogbo, Boluwaduro, Ifedayo, Ola-Oluwa, Aiyedade, Isokan,Ife-South and Ede north showed greater than 60% MDA participation. The average percentage of individual participation in MDA programs was found to be 56.09%, whereby, Ola-Oluwa LGA recorded the highest at 84.70% while the list MDA program participation was found at Ilesha East with 18.97%. Haematological findings indicated variable immune responses among patients with significant reduction in total WBC and lymphocyte counts, suggesting of immune suppression. Persistent monocytosis and reduced eosinophil levels were also observed.

As noted from WHO progress report on global programme to eliminate lymphatic filariasis, Pre-TAS is the first transmission assessment conducted in an intervention area to determine if transmission has been reduced to a level where it can be remain (or stay) even without control measures.^1^ To be eligible for Pre-TAS, in addition to 65% therapeutic coverage and 100% geographic coverage for at least 5 years, the intervention unit must have completed the fifth round of MDA at least 6 months prior to the assessment. Amanyi-Enegela et al. [18] reported that a pre-transmission assessment survey (pre-TAS) conducted in one sentinel site and one spot check site in Abaji and Kuje, Abuja, Nigeria in 2019 and was found to have LF antigenemia (LF Ag) < 2% (range 0.0% to 1.99%). In this work, the results obtained from the areas like Ola-Oluwa and Isokan, showed that high MDA rates likely contributed to effective suppression of parasite transmission.

The present results revealed that none of the LGAs had about 60% and above compliance in usage of LLINs, more especially in Orolu (28.71%), Oshogbo (27.35%), and Boluwaduro (31.39%), present a potential reinfection risk if MDA is discontinued prematurely. Richards et al., [19] highlighted the synergistic effect of MDA and vector control through LLINs in reducing the LF burden. Therefore, while drug uptake is encouraging, LLIN usage must be scaled up to sustain interruption of transmission.

The Abbott Filarial Test Strip (FTS) results in Table 3 showed a 0% prevalence of circulating filarial antigen (CFA) across all 7,388 individuals sampled across the 12 LGAs. This finding was well below the WHO threshold (<2%) for stopping MDA, indicating that LF transmission has been successfully interrupted in the surveyed areas. This outcome aligns with the WHO’s global LF elimination framework and mirrors successful outcomes reported in other endemic countries [16]. Biritwum et al., [20] reported a 0% CFA rate in Ghana after multiple MDA rounds, while Dorkenoo et al., [21] documented a similar outcome in Togo. Collectively, these findings reaffirm that consistent MDA campaigns and vector control can achieve the interruption of LF transmission, nevertheless, achieving 0% prevalence in pre-TAS does not imply the disease is eradicated. The impact of MDA is assessed through pre-transmission assessment surveys (Pre-TAS) and transmission assessment surveys which confirm if the prevalence of LF in an area has declined to levels at which MDA can be safely stopped [1]. Amanyi-Enegela et al., [22] reported that Transmission Assessment Surveys one (TAS 1) exercise was conducted in four distinct evaluation units (EUs) of two council areas within the FCT, Abuja, Nigeria. The TAS 1 exercise was carried out in Bwari and Gwagwalada evaluation units (EUs) due to successful achievement of pre-TAS thresholds indicating potential interruption of transmission. This correspond with the results obtained from the twelve (12) LGAs tested in this present study, suggesting for further progression to TAS 1 exercise in those 12 LGAs. The WHO emphasises the importance of post-MDA surveillance, continued for several years, to ensure no resurgence, particularly in communities with low LLIN usage or those at risk of reinfection due to migration from endemic regions [19].

Haematological analyses revealed statistically significant differences (p < 0.05) in immune cell counts between LF patients and healthy controls, particularly in WBCs, lymphocytes, monocytes, and eosinophils. Basophils were not found in all patient samples. Patients A and B exhibited marked decreases in WBCs, lymphocytes, neutrophils, and eosinophils, but an increase in monocyte levels (Figure 5). This pattern suggests parasite-induced immunosuppression, reflecting chronic antigen exposure and immune exhaustion, well-documented hallmarks of long-term filarial infection [23]. The reduction in eosinophils and neutrophils further indicates impaired adaptive and innate immune responses, while the elevated monocyte count may reflect compensatory activation of phagocytic and inflammatory pathways during chronic infection [12]. This persistent monocyte activity appeared to be associated with prolonged exposure to filarial antigens and may also contribute to the fibrotic and inflammatory tissue changes seen in LF-affected individuals.

In contrast, Patients C and D demonstrated elevated WBC, lymphocyte, neutrophil, and monocyte counts, suggesting an active immune response, possibly associated with antigenic stimulation, parasite death, or superimposed bacterial infection. The increased neutrophil levels imply ongoing inflammation, secondary infection, or acute tissue damage [24], while simultaneous lymphocytosis and monocytosis reflect an engaged adaptive and innate immune system. Such immune reactivation has been observed in individuals transitioning from immune tolerance to active inflammatory disease in lymphatic filariasis [25]. Whereas patient E demonstrated increased levels of WBCs, lymphocytes, and monocytes, but decreased neutrophils and eosinophils. This mixed immune profile suggests an immune imbalance, possibly driven by chronic immune regulation. The reduced granulocyte levels (neutrophils and eosinophils) may be attributed to the regulatory T cells (Treg) activity suppressing Th2-mediated responses, a mechanism frequently described in chronic lymphatic filariasis [23]. The suppression of eosinophils could reflect altered cytokine profiles, particularly reduced IL-5 levels, which is vital for eosinophil activation and survival [26]. This pattern likely represents an adaptive immune deviation in which Treg-mediated pathways dominate to limit excessive tissue-damage while maintaining low-grade immune surveillance during chronic infection.

Overall, the haematological patterns observed in this study demonstrated the heterogeneity of immune responses in LF patients, ranging from profound immunosuppression to hyper-inflammatory states. The haematological analyses of previously diagnosed LF patients revealed lingering immune alterations, characterised by variable levels of WBCs, lymphocytes, neutrophils, monocytes, eosinophils and the absence of basophils. The prevalence of monocytosis across most LF patients underscores the continued involvement of innate immunity, even in immunosuppressed states. These findings are consistent with reports of Mukherjee et al., [11] and Babu & Nutman [10] who described LF as a disease of immune dysregulation characterised by T-cell hypo-responsiveness, eosinophil dysfunction, and monocyte-driven chronic inflammation. Although the findings showed that Osun State is progressing toward complete LF elimination, post-MDA surveillance and strengthened community participation, particularly in improving LLIN usage and maintaining high MDA coverage, remain critical to sustain these gains and prevent resurgence.

## Data availability statement

The data of the original sample collections used in this study are included in the article/Supplementary Materials. Any other inquiries can be directed to the corresponding author.

## Ethics statement

This study was conducted in compliance with institutional and state ethical standards. The ethical clearance was obtained from the Ethical Review Committee of Adeleke University, Ede with the reference number given as AUERC/2025/IND/BCH/02, dated 18 September 2024 and Osun State Health Research Ethics Committee (OSHREC), Ministry of Health, Osun State, referenced as OSHREC/PRS/25/079 and dated 26 August 2024). All participants were fully informed of the study’s objectives, procedures, potential risks, and their rights. A written informed consent form was filled out by each participant before inclusion in the study. Confidentiality and anonymity were strictly upheld, and all participants retained the right to withdraw at any time without consequences. **Authors’ Contributions:** Conceptualisation, Project administration, Methodology, Data analysis, Writing original draft, Supervision and editing, N.S.C.; Project Supervision, review and editing, B.S.O., M.A.E., B.A.L., A.M.O.; Project administration, Supervision and data collection, B.A.L., A.T.A., A.M.O., A.T.A., B.S.O.; Conceptualisation, Writing review, data collection and data analysis, K.D.O., O.M.O., M.A.E., G.A.; Methodology, Project administration, project review and editing, O.O., O.D.A., I.A.A., O.E.O., O.O., U.E.; partly funding acquisition, H.A., B.A.L., U.E., I.A.A.;

## Acknowledgements

The authors appreciate the Department of Public Health in the Ministry of Health, Osun State, Nigeria for providing the logistics and necessary support during the pre-transmission survey. Appreciation to the Department of Biochemistry, Adeleke University, Ede, Osun State, Nigeria for spearheading and providing the enabling support for the success of this research work. Moreover, thanks to the Bio-repository and clinical virology laboratory unit of University College Hospital, Ibadan, Oyo State, Nigeria, for providing the space and equipment for certain analyses. Thanks to University of Osun Teaching Hospital, Osogbo, Osun State, Nigeria for making their specialists available to perform the painstaking sample collection from the patients.

